# Immune responses to a single dose of the AZD1222/Covishield vaccine at 16 weeks in individuals in Sri Lanka

**DOI:** 10.1101/2021.07.26.21261122

**Authors:** Chandima Jeewandara, Dinuka Guruge, Pradeep Darshana Pushpakumara, Achala Kamaladasa, Inoka Sepali Aberathna, Shyrar Tanussiya, B Banuri Gunasekera, Ayesha Wijesinghe, Osanda Dissanayake, Heshan Kuruppu, Thushali Ranasinghe, Deshni Jayathilaka, Shashika Dayarathna, Dinithi Ekanayake, MPDJ Jayamali, Nayanathara Gamalath, Anushika Mudumkotuwa, Gayasha Somathilake, Madhushika Dissanayake, Michael Harvie, Thashmi Nimasha, Deshan Madusanka, Tibutius Jayadas, Ruwan Wijayamuni, Lisa Schimanski, Pramila Rijal, Tiong .K. Tan, Alain Townsend, Graham S. Ogg, Gathsaurie Neelika Malavige

## Abstract

**Introduction:** Due to limited access to vaccines, many countries have only administered a single dose of the AZD1222, while the dosage intervals have increased ≥ weeks. We sought to investigate the immunogenicity of a single dose of vaccine at ≥ 16 weeks.

**Methods:** SARS-CoV-2 specific antibodies in 553 individuals and antibodies to the receptor binding domain (RBD) of the Wuhan virus (WT) and the variants of concern (VOCs), ACE2 receptor blocking antibodies, ex vivo and cultured IFNγ T cell responses and B cell ELISpot responses were investigated in a sub-cohort.

**Results:** The seropositivity rates in those >70 years of age (93.7%) was not significantly different compared to other age groups (97.7 to 98.2, Pearson Chi-Square = 7.8, p-value = 0.05). The antibody titres (antibody index) significantly declined (p<0.0001) with increase in age. 18/69 (26.1%) of individuals did not have ACE2 receptor blocking antibodies, while responses to the RBD of WT (p=0.03), B.1.1.7 (p=0.04) and B.1.617.2 (p=0.02) were significantly lower in those who were >60 years. Ex vivo IFN γ T cell ELISpot responses were seen in 10/66 (15.1%), while only a few expressed CD107a. However, >85% had a high frequency of cultured IFNγ T cell ELISpot responses and B cell ELISpots.

**Conclusion:** Virus specific antibodies were maintained at ≥ 16 weeks after receiving a single dose of AZD1222, although levels were lower to VOCs, especially in older individuals. A single dose induced a high frequency of memory T and B cell responses.

## Introduction

Although there were high expectations that the COVID-19 pandemic will be controlled in 2021, the number of cases and deaths due to COVID-19 continues to increase, with infections predominantly seen in lower middle income (LMICs) and low income countries(Medicine, 2021). While higher income countries have vaccinated at least over 30% of their population as of early July 2021, many LMICs and low-income countries have administered one dose of a COVID-19 vaccine to <5% of their population (Hannah Ritchie, 2021). Many countries increased the interval between the two doses, in order to vaccinate as many individuals as possible, with a single dose (Pimenta et al., 2021).

The AZD1222 vaccine is a chimpanzee adenovirus vector vaccine, containing the spike protein of SARS-CoV-2 virus (Voysey et al., 2021a). Although in initial clinical trials, the interval between the two doses were 28 days (Ramasamy et al., 2021), later studies showed that increasing the dose interval to twelve weeks, increased the efficacy of the vaccine to 81.3% compared to an efficacy rate of 55.1% when the interval between the two doses was <6 weeks(Voysey et al., 2021b). However, due to the sharp increase in the number of cases, some countries such as India and Canada, increased the gap between the doses to 16 weeks, in order to vaccinate a larger proportion of the population with a single dose (Corte, 2021; Tauh. T, 2021). The COVID-19 vaccination in Sri Lanka commenced with immunizing the health care workers with AZD1222/Covishield vaccine, which was later rolled out to the public in the Colombo district. However, due to shortfalls in a continuous supply, Sri Lanka did not receive sufficient doses to give the second dose as planned at 12 weeks since receiving the first dose of the vaccine. Therefore, there is currently a large population, which has received the first dose 16 weeks ago, awaiting the second dose of the vaccine.

It was recently shown that the antibodies induced by a single dose of AZD1222 persisted for a year, but the levels on day 180, were half the levels seen on day 28 post-vaccination (Flaxman, 2021). However, a single dose of the vaccine was shown to be only 33% effective against the delta variant, which is the predominant variant currently, seen in many countries (Iacobucci, 2021). Many countries, which gave a single dose of AZD1222 to their population but are unable to secure the second dose in time, with the delta variant emerging are in a dilemma. As Sri Lanka faces a similar situation, we sought to investigate the antibody and T cell responses, including memory T cell and memory B cell responses in individuals of varying age who received the vaccine ≥ 16 weeks ago.

## Methods

### Study participants

553 individuals, who received their first dose of the AZD1222/Covishield vaccine between the 15^th^ of February to 4^th^ March 2021, were included in the study following informed written consent (16 to 17 weeks since receiving the first dose). Demographic and the presence of comorbidities such as diabetes, hypertension, cardiovascular disease and chronic kidney disease was determined by a self-administered questionnaire at the time of recruitment. Blood samples were obtained from all individuals to determine seropositivity rates at ≥16 weeks since obtaining the first dose of the vaccine, while T cell studies, ACE2 blocking and variant specific antibody responses for SARS-CoV-2 were carried out in only 69 individuals, who were randomly selected from different age categories. Ethics approval was obtained from the Ethics Review Committee of University of Sri Jayewardenepura.

### Detection of SARS-CoV-2 specific antibodies

The presence of SARS-COV-2 specific antibodies were detected by using the Wantai SARS-CoV-2 Ab ELISA (Beijing Wantai Biological Pharmacy Enterprise, China), which detects IgM, IgG and IgA antibodies to the receptor binding domain (RBD) of the spike protein. A cut-off value for each ELISA was calculated according to manufacturer’s instructions. Based on the cut off value, the antibody index was calculated by dividing the absorbance of each sample by the cutoff value, according to the manufacturer’s instructions.

### Surrogate neutralizing antibody test (sVNT) to detect ACE2 receptor blocking antibodies and RBD binding antibodies detected by HAT

The surrogate virus neutralization test (sVNT) (Tan et al., 2020), which measures the percentage of inhibition of binding of the RBD of the S protein to recombinant ACE2 (Tan et al., 2020) (Genscript Biotech, USA) was carried out according the manufacturer’s instructions as previously described by us (C. Jeewandara et al., 2021). Inhibition percentage ≥ 25% in a sample was considered as positive for ACE2 receptor blocking antibodies.

In order to quantify IgG antibodies to the RBD, WANTAI SARS-CoV-2 quantitative IgG ELISA (CAT-WS-1396) was used. The assay was carried out and interpreted according to the manufacturer’s instructions.

### Haemagglutination test (HAT) to detect antibodies to the receptor binding domain (RBD)

The HAT was carried out as previously described using the B.1.1.7 (N501Y), B.1.351 (N501Y, E484K, K417N) and B.1.617.2 versions of the IH4-RBD reagents (Townsend et al., 2020), which included the relevant amino acid changes introduced by site directed mutagenesis. The assays were carried out and interpreted as previously described (Chandima Jeewandara et al., 2021). The HAT titration was performed using 7 doubling dilutions of serum from 1:20 to 1:1280, to determine presence of RBD-specific antibodies. The RBD-specific antibody titre for the serum sample was defined by the last well in which the complete absence of “teardrop” formation was observed. A titre of 1:20 was considered as a positive response, as previously determined by us (Kamaladasa et al., 2021).

### Ex vivo and cultures IFNγ ELISpot assays

Ex vivo IFNγ ELISpot assays were carried out using freshly isolated peripheral blood mononuclear cells (PBMC) obtained from 66 individuals. Due to the limitation in the number of cells, cultured ELISpots were only carried out in 13 individuals. Individuals for T cell assays were randomly recruited from the study participants, and we included those who consented to provide an additional blood volume for T cell assays (7ml), in addition to the antibody assays (5ml). For ex vivo ELISpot assays, two pools of overlapping peptides named S1 (peptide 1 to 130) and S2 (peptide 131 to 253) covering the whole spike protein (253 overlapping peptides) were added at a final concentration of 10 µM and incubated overnight as previously described (Malavige et al., 2008; Peng et al., 2020). For cultured ELISpot assays, 5.0×10^6^ PBMCs were incubated for 10 days with 200µl of 40µM peptide pool of overlapping S peptides in a 24 well plate. IL-2 was added on day 3 and 7 at a concentration of 100units/ml. All cell lines were routinely maintained in RPMI 1640 supplemented with 2mM L-glutamine, 100IU/ml penicillin and 100µg/ml plus 10% human serum at 37°C, in 5% CO_2_ (Jeewandara et al., 2018).

All peptide sequences were derived from the wild-type consensus and were tested in duplicate. For ex vivo ELISpot assays, 100,000 cells/well were added and for cultures ELISpots, 50,000 cells/well were added and PHA was included as a positive control of cytokine stimulation and media alone was applied to the PBMCs as a negative control. Briefly, ELISpot plates (Millipore Corp., Bedford, USA) were coated with anti-human IFNγ antibody overnight (Mabtech, Sweden). The plates were incubated overnight at 37°C and 5% CO_2_. The cells were removed, and the plates developed with a second biotinylated antibody to human IFNγ and washed a further six times. The plates were developed with streptavidin-alkaline phosphatase (Mabtech AB) and colorimetric substrate, the spots were enumerated using an automated ELISpot reader (AID Germany). Background (PBMCs plus media alone) was subtracted and data expressed as number of spot-forming units (SFU) per 10^6^ PBMCs. A positive response was defined as mean±2 SD of the background responses.

### Intracellular cytokine staining

The expression of CD107a and IFN-g were determined on both CD4 and CD8 T cells by flowcytometry in freshly isolated PBMC as described previously (Wijeratne et al., 2019). Briefly, PBMCs were incubated with CD107a FITC (Biolegend, USA) for 30□minutes in RPMI 1640 and 10% heat inactivated human serum (Sigma Andrich). Cells were stimulated with overlapping peptides pool of SARS-CoV-2 spike protein for 2 hours at 1mM concertation before adding monensin (Biolegend, USA). The PBMC were incubated for a further 14 hours before staining with with anti□CD3 APC Cy7 (clone OKT3), anti□CD8 BV650 (clone SK1) and anti-CD4 PB (clone OKT4). Then the cells were fixed with fixation buffer and permeabilized with perm wash buffer (Biolegend, USA) and stained for IFN-γ APC (clone 4S. B3). Live/Dead fixable aqua dead cell stain (Thermo Fisher Scientific, USA) was used according to the manufacturer’s protocol to exclude dead cells. Cells were acquired on a BD FACSAria III Cell Sorter using DIVA v8 software (BD Biosciences, USA). For each donor, unstimulated cells were included as a negative control. Flow cytometry data were analyzed using FlowJo v.10.7.1 software (FlowJo). Fluoresces Minus One (FMO) controls were used to draw the gates for both CD107a and IFN-γ (supplementary figure 1).

**Figure 1:**
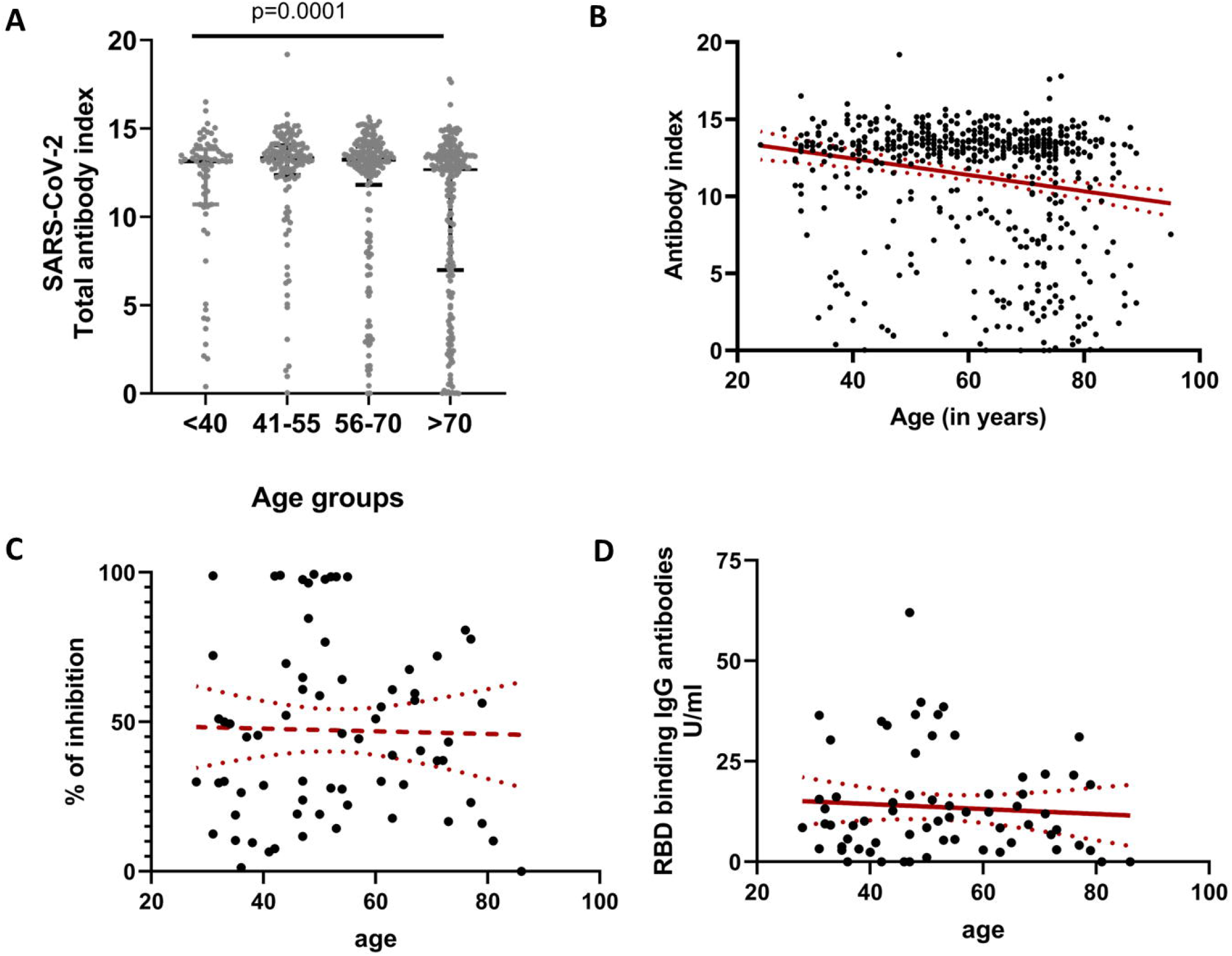
SARS-CoV-2 specific antibodies in individuals of different age groups who have received a single dose of AZD1122, ≥16 weeks ago. The SARS-CoV-2 specific total antibodies (IgG, IgA and IgM) were measured in those who were <40 (n=56), 41 to 55 years (n=132), 56 to 70 (n=171) and in those >70 years of age (n=205), by the Wantai total antibody assay. The Kruskal-Wallis test (two-tailed) was used to determine the differences in antibody levels between the different age groups (A). The age was correlated with total antibody levels (Spearman’s r=-0.18, p<0.0001) (B), ACE2 receptor blocking antibodies (Spearman’s r=0.02, p=0.81) in a subset of individuals (n=69) (C) and with antibodies to the RBD detected by HAT (Spearman’s r=-0.16, p=0.89) (D). All tests were two sided. The error bars indicate the median and the interquartile ranges in (A) and the 95% CI in other figures.

### B cell ELISpot assays

The frequency of SARS-CoV-2 spike and N protein specific memory B cells were assessed using B cell ELISpot assays. Briefly, freshly isolated PBMCs were stimulated in a 24 well plate using IL-2 and R848 (a TLR 7/8 agonist) in RPMI supplemented with 10% fetal bovine serum, 1% penicillin streptomycin and 1% glutamine at 4 million cells/well and incubated at 37 °C with 5% CO_2_ for 3 days. They were then washed and rested overnight and 100,000 cells/well were added. 50,000 cells/well were added to the positive control wells. A Human IgG ELISpot kit (Mabtech 3850-2A) was used according to the manufacturer’s instructions to quantify IgG-secreting cells specific to SARS-COV2 S1, S2 and N recombinant proteins, which were coated at 2µg/ml in phosphate buffered saline (PBS). All experiments were carried out in duplicate and anti-human IgG monoclonal capture antibodies, was used as a positive control, and media alone as a negative control. The spots were enumerated using an automated ELISpot reader (AID Germany).

### Statistical analysis

The 95% confidence intervals for seropositivity for each age category were calculated using the R software (version 4.0.3) and R-studio (version 1.4.1106). Pearson Chi Square Association tests were performed at a confidence level of 95% using the R software to identify the statistically significant associations of the age categories and the sex of the individuals with seropositivity. Kruskal-Wallis test was used to determine the differences between the levels of antibodies between different age groups. Spearman’s correlation coefficient was used to determine the correlation between antibody, T cell responses and the age of an individual.

## Results

The median age of the study participants was 64 years (range 24 to 95 years). At ≥16 weeks, 553/573 (96.5%) individuals were seropositive for SARS-CoV-2. The seropositivity and the number of individuals in each age group is shown in table 1. Although the seropositivity rates were lower in those >70 years of age (93.7%) compared to other age groups (97.7 to 98.2%) this was not significant (Pearson Chi-Square = 7.8, p-value = 0.05). However, the SARS-CoV-2 total antibody index values (an indirect measure of the antibody titres), was significantly different (p=0.0001) in these four groups (Figure 1A). There was no difference in the total antibody levels (antibody index) in age groups <40, 41 to 55 and 56 to 69, although the antibody levels of those >70 years of age was significantly lower than the other groups. The antibody titres (antibody index) significantly declined (p<0.0001) with increase in age (Figure 1B).

**Table 1:**
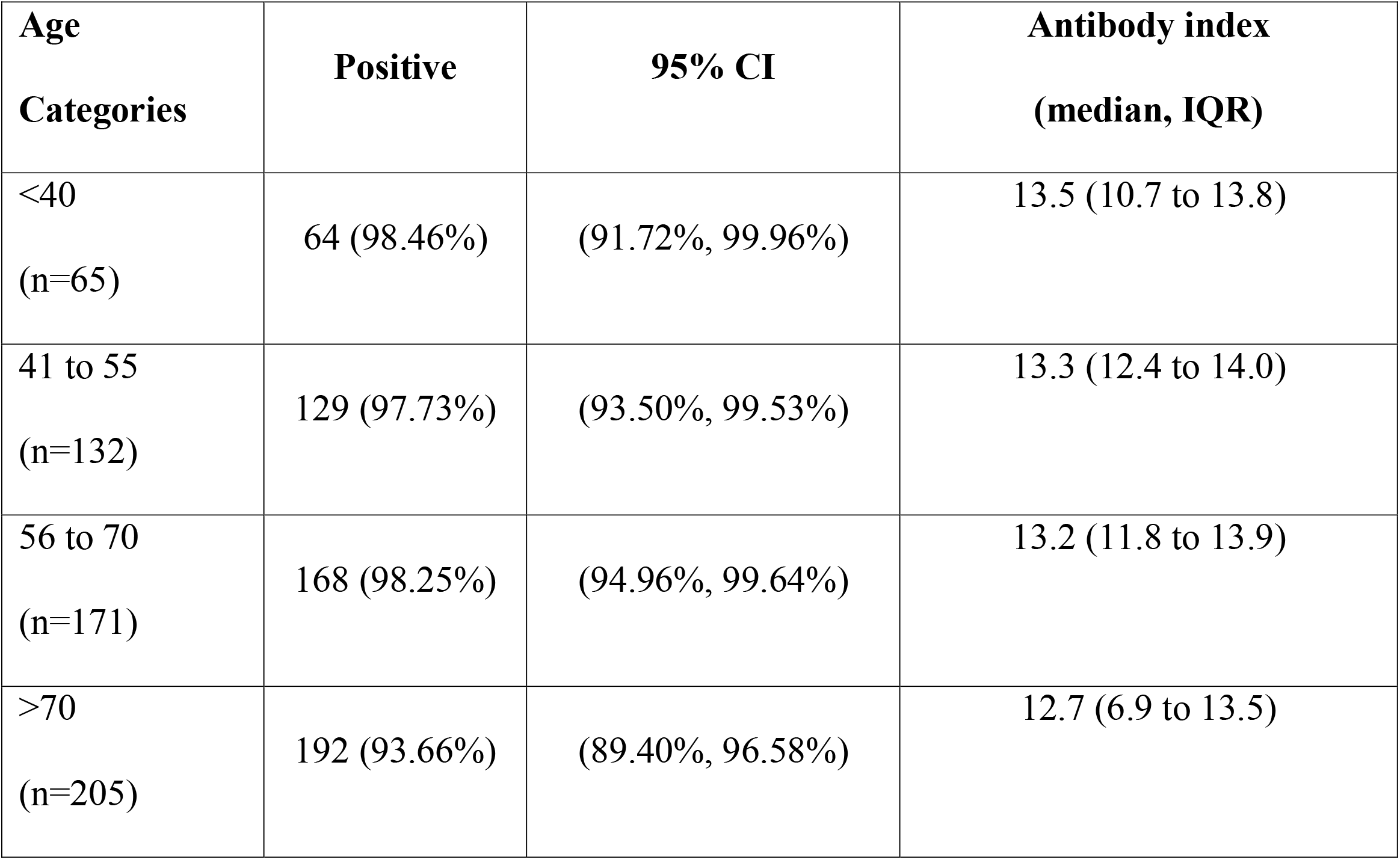
Age categories and SARS-CoV-2 antibody positivity in individuals who received a single dose of the AZD1222 vaccine ≥ 16 weeks.

301/573 (52.5%) individuals were females and there was no difference (Pearson Chi-Square = 0.46, p-value = 0.5) in the seropositivity in females (97.01%, 95% CI 94.4% to 98.6%) compared to males (95.9%, 95% CI 92.9% to 97.9%).

### Presence of ACE2 receptor blocking antibodies and RBD binding IgG antibodies

We used the surrogate neutralizing test (sVNT) to determine the presence of ACE2 receptor blocking antibodies in a sub cohort of the above individuals (n=69). A percentage inhibition of 25% was considered as a positive result as previously described (C. Jeewandara et al., 2021). 51/69 (73.9 %) of individuals had ACE2 receptor blocking antibodies above the cut-off value. There was correlation (Spearmans r=0.02, p=0.81) between the age and ACE2 receptor blocking antibodies (Figure 1C). We also measured IgG antibodies to the RBD in these individuals, and again there was no correlation with the age (Spearman’s r=0.01, p=0.89) (Figure 1D). However, 5 individuals had undetectable levels of IgG antibodies to the RBD.

### Antibodies to the receptor binding domain of the wild type and SARS-CoV-2 variants

Antibodies to the RBD were measured by HAT to the wilt type (WT), B.1.1.7, B.1.351 and B.1.617.2 in 69 individuals ranging from 20 to 85 years of age. There was no difference in the HAT titres to the B.1.1.7 (p>0.99) and B.1.617.2 (p=0.90) compared to HAT titres to the WT, whereas the titres to the B.1.351 was significantly lower (p<0.0001) (Figure 2A). A HAT titre of 1:20 was considered as a positive result to the RBD by the HAT assay, as previously described(Kamaladasa et al., 2021). Of these 69 individuals, 18/69 (26.1%) had a positive response to the WT, 19/69 to B.1.1.7, 3/69 to B.1.351 and 18/69 to B.1.617.2.

**Figure 2:**
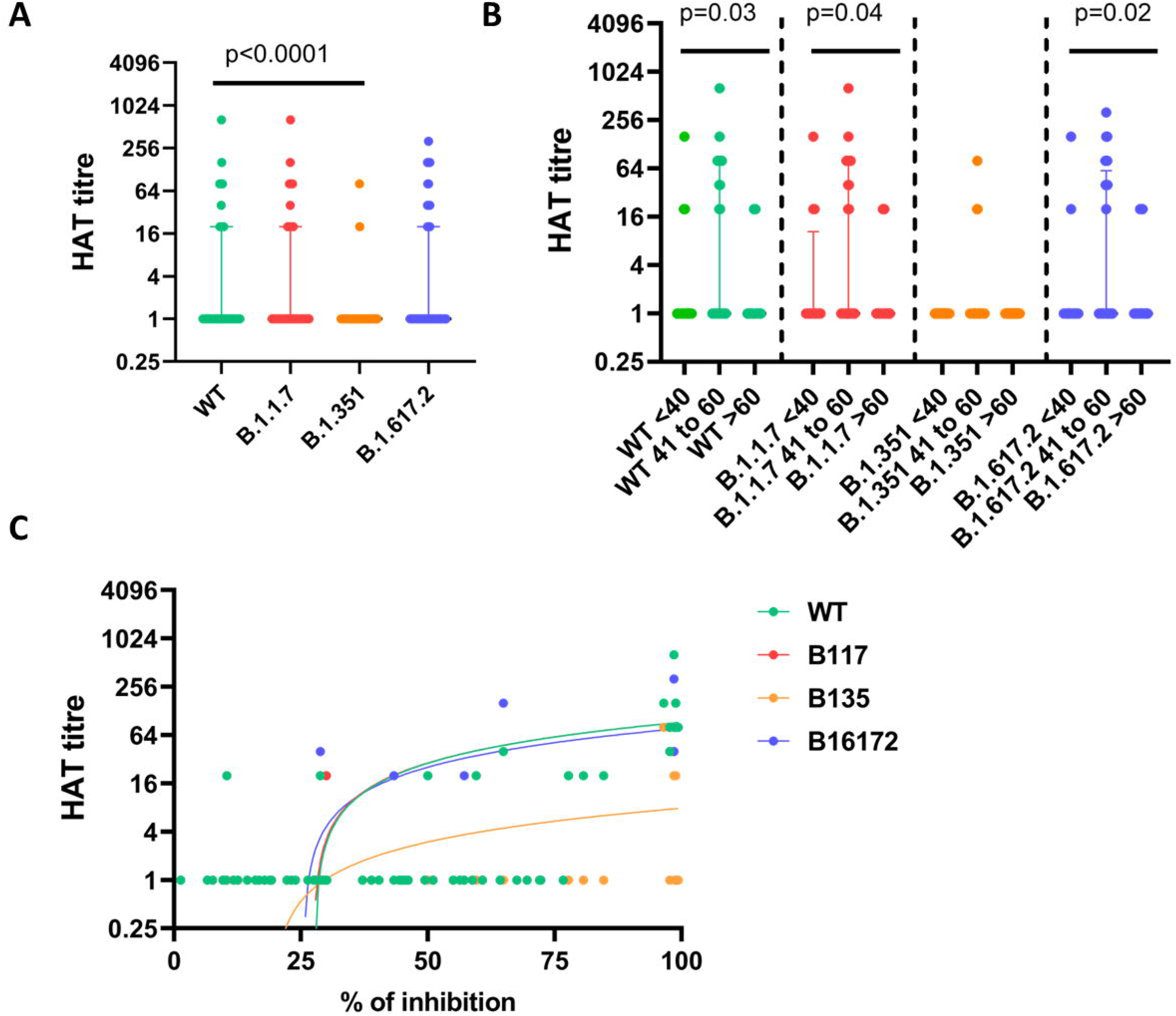
Antibody responses to the SAR-CoV-2 Wuhan, wild type variant and other variants of concern. Antibody responses to the RBD of the wild type (WT), B.1.1.7, B.1.351 and B.1.617.2 was determined by HAT in 69 individuals (A) and also in these cohort of individuals, when categorized into three age groups (B). The antibody responses for the variants were correlated with the ACE2 receptor blocking antibody levels, which significantly and positively correlated with the HAT titres for WT (Spearman’s r=0.62, p<0.0001), B.1.1.7 (Spearman’s r=0.61, p<0.0001), B.1.351 (Spearman’s r=0.30, p=0.01) and B.1.617.2 (Spearman’s r=0.64, p<0.0001) (C). All tests were two sided. The error bars indicate the median and the interquartile ranges.

Of the 69 individuals, 17 were between the ages of 20 to 40, 30 individuals between the ages of 41 to 60 and 22 individuals >60 years old. The antibody responses to WT (p=0.03), B.1.1.7 (p=0.04) and B.1.617.2 (p=0.02) were significantly lower in those who were >60 years, compared to other groups, where there was no difference in responses to B.1.351 (Figure 2B). Of those who were >60 years, 3/22 (13.6%) had a positive response to WT, 3 for B.1.1.7, 4/22 to B.1.617.2 and none for B.1.351. The ACE2 receptor blocking antibody levels significantly and positively correlated with the HAT titres for WT (Spearman’s r=0.62, p<0.0001), B.1.1.7 (Spearman’s r=0.61, p<0.0001), B.1.351 (Spearman’s r=0.30, p=0.01) and B.1.617.2 (Spearman’s r=0.64, p<0.0001) (Figure 2C).

### Ex vivo and cultured IFNγ T cell ELISpot responses

Ex vivo IFNγ T cell ELISpot assays were carried out in 66 individuals (same cohort of individuals used in sVNT and HAT assays) for 253 overlapping peptides, divided in to two peptide pools, S1 (peptide 1 to 130) and S2 (peptide 131 to 253). A positive IFNγ ELISpot response was defined as mean±2 SD of the background responses and accordingly, a positive cut-off threshold was set as 200 spot forming units (SFUs)/1 million PBMCs. 10/66 (15.1%) individuals responded to S1 and 5/66 (7.6%) to S2 (Figure 3A). However, there was no correlation between the age of individuals and the frequency of the ex vivo ELISpot responses for S1 (Spearman’s r=-0.06, p=0.59), S2 (Spearman’s r=0.14, p=0.29) or S (Spearman’s r=-0.00, p=0.94) (Figure 3B).

**Figure 3:**
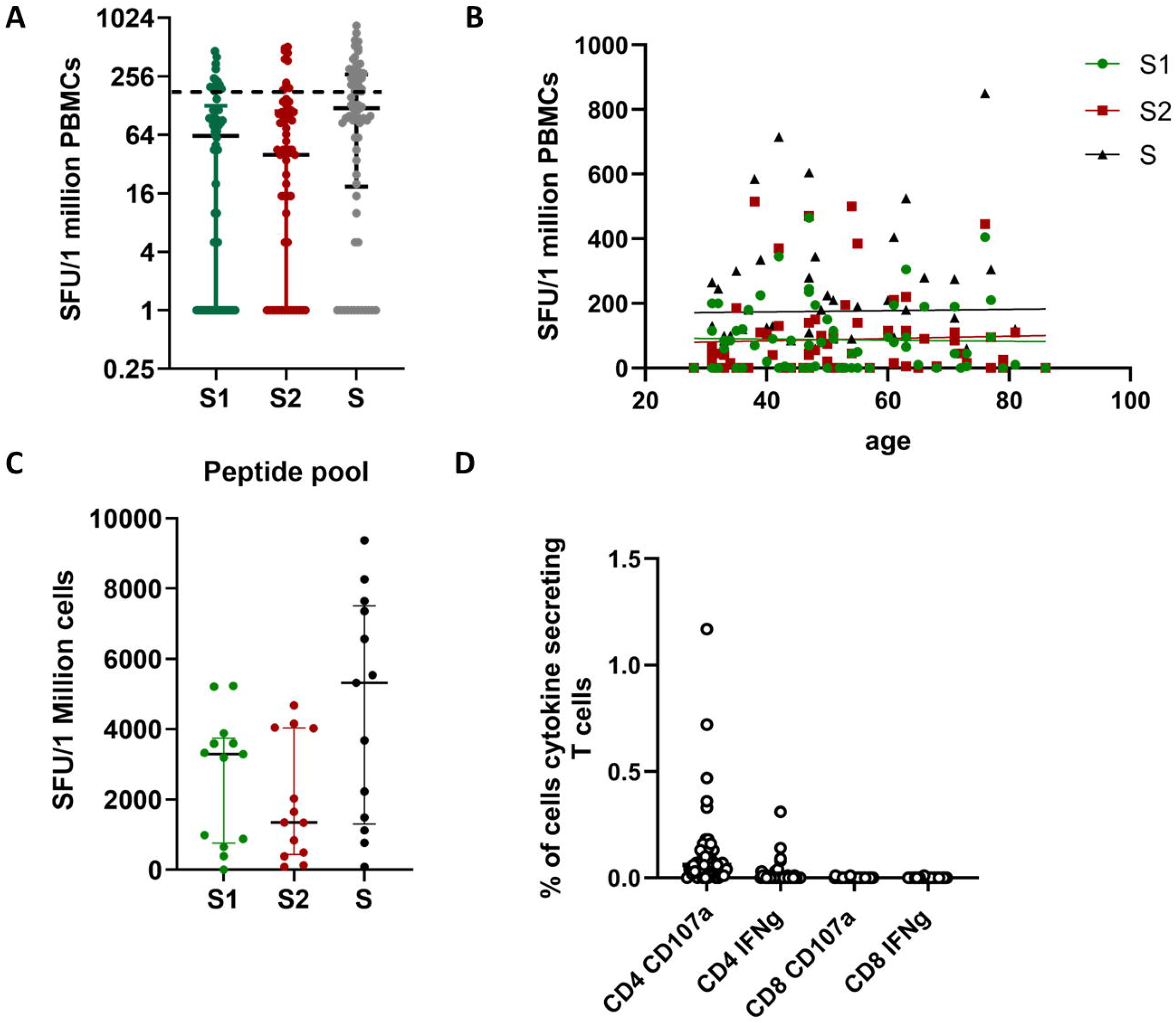
T cell responses to the SARS-CoV-2 spike protein overlapping peptides in individuals who have received a single dose of AZD1122, ≥ 16 weeks ago. The frequency of S protein specific ex vivo IFNγ ELISpot responses to two over lapping peptide pools (S1 and S2) were assessed (n=16) (A) and was also correlated with age for S1 (Spearman’s r=-0.06, p=0.59), S2 (Spearman’s r=0.14, p=0.29) or S (Spearman’s r=-0.00, p=0.94) (B). The cultured ELISpot IFNγ ELISpot responses were assessed (n=13), for S1 and S2 (C) and the intracellular cytokine responses (CD107a and IFNγ) to the whole spike overlapping pool of peptides for S were also assessed (n=46). All tests were two sided. The error bars indicate the median and the interquartile ranges.

Due to limitation in cell numbers, cultured ELISpot assays were only possible in 13 individuals. A positive IFNγ ELISpot response was defined as mean±2 SD of the background responses and accordingly, a positive cut-off threshold was set at 83 SFUs/1 million cells. Except for one individual, 12/13 individuals had robust T cell responses when assessed after short term culture of T cells. As seen with ex vivo ELISpot responses there was no significant difference (p=0.27) between responses to S1 pool compared to S2 pool of peptides (Figure 3C).

### Intracellular cytokine responses

We also carried out intracellular cytokine staining to explore the frequency of S overlapping peptide-specific CD4+ and CD8+ T cells that are able to produce IFNγ or degranulated (CD107a). Due to the limitation in the number of cells, these assays were carried out in only 46 individuals of the 66 individuals in whom ELISpot assays were performed. None of the individuals had any IFNγ producing S protein specific D8+ T cells, although two individuals had a low frequency of CD107a expression. In contrast, except for 2 individuals, the S protein specific CD4+ T cells of 44/46 individuals, expressed varying levels of CD107a, while 12/46 individuals had detectable IFNγ -producing T cells (Figure 3D).

### B cell ELISpot responses to investigate antigen secreting B cells

B cell ELISpots were carried out in 40 individuals as there were limited number of cells to carry out B cell ELISpots in all 66 individuals, included in ex vivo IFNγ ELISpot assays. As a positive response was defined as mean±2 SD of the background responses and accordingly, a positive cut-off threshold was set at 39.3 antigen secreting cells (ASCs)/1 million for the S1 protein, 27.7 for S2 and 16.0 for N recombinant protein. Accordingly, 36/40 (90%) had responses to S1, 35/40 (87.5%) for S2 and 30/45 (66.7) for N protein. However, the S1 responses (median 90, IQR 46.25 to 130 ASCs/million cells) were significantly higher (p=0.04) than for S2 (median 70, IQR 46.25 to 87.5 ASCs/million cells) and N (p<0.0001, median 50, IQR 25 to 73.75 ASCs/million cells) (Figure 4A). Interestingly, the S1 (Spearmans’ r=0.51, p=0.0008), S2 (Spearmans’ r=0.46, p=0.003) and N (Spearmans’ r=0.41, p=0.008), significantly and positively correlated with age (Figure 4B).

**Figure 4:**
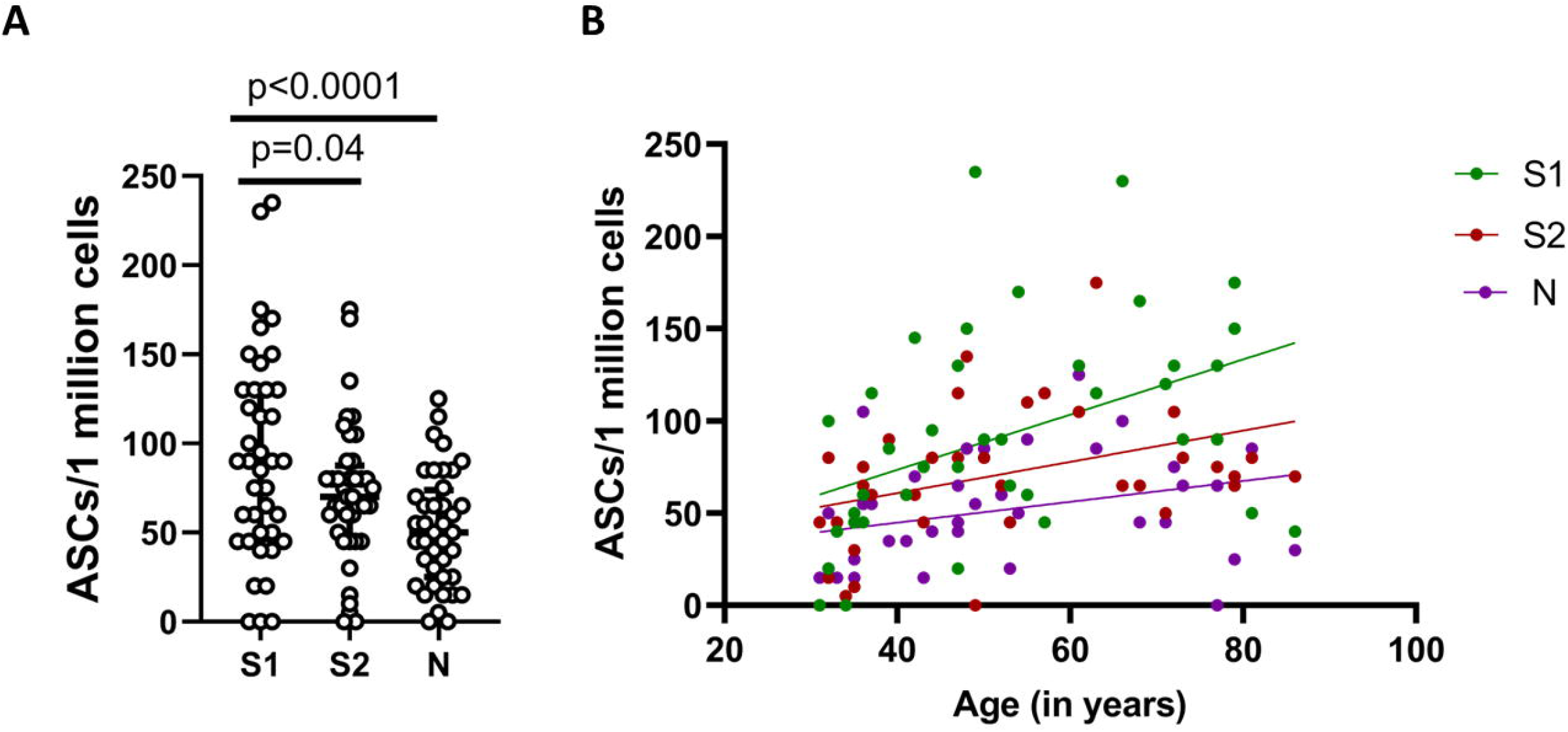
B cell ELISpot responses to the SARS-CoV-2 S1, S2 and N proteins in individuals who have received a single dose of AZD1122, ≥16 weeks ago. B cell ELISpots were carried out for S1, S2 and N2 recombinant protein in individuals (n=40), ≥16 weeks after receiving the vaccine to determine the antibody secreting cells (ASCs) (A). The frequency of ASCs S1 (Spearmans’ r=0.51, p=0.0008), S2 (Spearmans’ r=0.46, p=0.003) and N (Spearmans’ r=0.41, p=0.008) significantly and positively correlated with age (B). The Wilcoxon test was used to determine the differences between the frequency of ASCs for S1, S2 and N. All tests were two sided. The error bars indicate the median and the interquartile ranges.

## Discussion

Between 16 to 17 weeks since receiving the first dose of AZD1222, 93.6% to 98.5% were seropositive for SARS-CoV-2. Although the total antibody levels waned with age, the total antibody levels were similar to the levels seen by us at 4 weeks following a single dose of the vaccine in health care workers in Sri Lanka (Chandima Jeewandara et al., 2021). However, by 16 weeks, 26.1% did not have ACE2 receptor blocking antibodies and ∼74% of individuals did not have antibodies to the RBD when assessed by the HAT assay to the WT. Importantly, those who were >60 years of age, only 13.6% gave a positive response against antibodies to the RBD to WT, B.1.1.7 and B.1.617.2, whereas no one had antibodies to B.1.351. Therefore, although total antibodies to SARS-CoV-2 appear to persist unchanged from 4 to 16 weeks, ACE2 receptor blocking antibodies appear to decline especially in older individuals, thereby possibly resulting in reduced protection.

In this context of sixteen weeks post a single vaccination the HAT test was considerably less sensitive (26.1%) than the Wantai SARS-CoV-2 Ab ELISA (96.5%) or ACE2 Blocking assay (73.9%). This contrasts with previous results after mild infection where the HAT was slightly more sensitive than the ACE2 blocking assay (Kamaladasa et al., 2021), and serum donors post symptomatic illness, where HAT and ACE2 blocking assays performed similarly and correlated well with neutralising titre (Lamikanra et al in Press Transfusion). All of the assays described here measure antibodies to the isolated Receptor Binding Domain, so these variations suggest qualitative differences in the antibodies in these different contexts. HAT depends on antibodies that can agglutinate red cells so may be more sensitive in situations where IgM persists, although the majority of IgG monoclonal antibodies are also capable of agglutinating red cells(Townsend et al., 2020) (Townsend ref). Fourteen to twenty one days days after a single dose the AZ vaccine is ∼67% effective against symptomatic infection with the alpha variant (Nasreen et al., 2021). This suggests that a positive HAT test at 1:20 in this context may be detecting a level of antibody above what is needed for this level of protection.

Apart from antibodies, T cell responses have been shown to correlate with protection against severe COVID-19 illness (Kalimuddin et al., 2021; Sadarangani et al., 2021). SARS-CoV-2 spike protein specific T cells were seen very early following the first dose of the vaccine, before the appearance of neutralizing antibodies, and it is speculated that T cells in the absence of neutralizing antibodies may be protective (Kalimuddin et al., 2021). In our cohort, this represents ex vivo 7.6% to 15.1% of individuals. This is consistent with other studies, where it was shown that ex vivo IFNγ T cell responses detected after a single dose of a mRNA vaccine, became undetectable after 21 days, but robust T cell responses were seen after T cell expansion in vitro(Kalimuddin et al., 2021). We too observed that all 12/13 individuals, had robust memory T cell responses after in vitro stimulation. IFNγ production by CD8+ T cells were also not detected by intracellular cytokine staining, although spike protein specific CD4+ T cells demonstrated variable levels of CD107a expression and IFNγ production. It has been shown that the CD4+ T cell responses were of a higher magnitude than the CD8+ T cell responses to spike protein overlapping peptides following vaccination with the mRNA vaccines and AZD1222 (Ewer et al., 2021; Woldemeskel et al., 2021). Therefore, our results show that the CD4+ T cell responses dominate the spike protein specific T cell response at 16 weeks, following vaccination.

Apart from antibody responses, memory B cells have shown to be induced by infection and following vaccination and have been shown to be generated in patients with acute COVID-19 (Byazrova et al., 2021). Such memory B cells have shown to be detectable many months following natural COVID-19 infection(Turner et al., 2021). Memory B cells following COVID-19 vaccines have not been extensively studied and we found that 36/40 (90%) of individuals had a high frequency of responses to the S1 subunit of the spike protein, 16 weeks after a single dose of AZD1222. Interestingly, a large proportion of individuals responded to the N protein as well, although the responses were of low frequency. This could be due to the cross reactivity of B cell responses of N proteins of other seasonal coronaviruses, as the N protein is highly conserved (Beretta et al., 2020; Nguyen-Contant et al., 2020). However, a few individuals did have a high frequency of ASCs to the N protein, which could have been due to possible asymptomatic infection, that could have occurred during this time period. Interestingly, the frequency of ASC B cell responses significantly correlated with age, suggesting that, seasonal coronavirus induced memory B cell responses could be boosted by the vaccine, in older individuals.

In summary, we show that ≥ 16 weeks from receiving a single dose of the AZD1222 vaccine, the antibody responses to the SARS-CoV-2 were maintained, although the levels were lower in older individuals. Although the antibodies to the RBD of VOCs, were not detected or were very low in most individuals, the majority had a high frequency of memory T and B cell responses.

## Data Availability

All data is available within the manuscript, figures and the supplementary figures.

## Data Availability

All data is available within the manuscript, figures and the supplementary figures.

## Acknowledgement

We are grateful to the World Health Organization, UK Medical Research Council and the Foreign and Commonwealth Office for support. T.K.T. is funded by the Townsend-Jeantet Prize Charitable Trust (charity number 1011770) and the EPA Cephalosporin Early Career Researcher Fund. A.T. are funded by the Chinese Academy of Medical Sciences (CAMS) Innovation Fund for Medical Science (CIFMS), China (grant no. 2018-I2M-2-002). We thank WBP, CBP, EGB and ANB for generous donations to the TJP Trust to support the production and distribution of HAT reagents.

## References

Beretta, A., Cranage, M., Zipeto, D., 2020. Is Cross-Reactive Immunity Triggering COVID-19 Immunopathogenesis? Frontiers in immunology 11, 567710.

Byazrova, M., Yusubalieva, G., Spiridonova, A., Efimov, G., Mazurov, D., Baranov, K., Baklaushev, V., Filatov, A., 2021. Pattern of circulating SARS-CoV-2-specific antibody-secreting and memory B-cell generation in patients with acute COVID-19. Clin Transl Immunology 10, e1245.

Corte, M.F., 2021. Delayed second dose is a winning strategy for vaccine-starved countries like India, The Econimic Times. The Economic Times India, India.

Ewer, K.J., Barrett, J.R., Belij-Rammerstorfer, S., Sharpe, H., Makinson, R., Morter, R., Flaxman, A., Wright, D., Bellamy, D., Bittaye, M., Dold, C., Provine, N.M., Aboagye, J., Fowler, J., Silk, S.E., Alderson, J., Aley, P.K., Angus, B., Berrie, E., Bibi, S., Cicconi, P., Clutterbuck, E.A., Chelysheva, I., Folegatti, P.M., Fuskova, M., Green, C.M., Jenkin, D., Kerridge, S., Lawrie, A., Minassian, A.M., Moore, M., Mujadidi, Y., Plested, E., Poulton, I., Ramasamy, M.N., Robinson, H., Song, R., Snape, M.D., Tarrant, R., Voysey, M., Watson, M.E.E., Douglas, A.D., Hill, A.V.S., Gilbert, S.C., Pollard, A.J., Lambe, T., Oxford, C.V.T.G., 2021. T cell and antibody responses induced by a single dose of ChAdOx1 nCoV-19 (AZD1222) vaccine in a phase 1/2 clinical trial. Nature medicine 27, 270–278.

Flaxman, A.a.M. Natalie, and Jenkin, Daniel and Aboagye, Jeremy and Aley Parvinder K. and Angus Brian John and Belij-Rammerstorfer, Sandra and Bibi, Sagida and Bittaye, Mustapha and Cappuccini, Federica and Cicconi, Paola and Clutterbuck, Elizabeth and Davies, Sophie and Dejnirattisai, Wanwisa and Dold, Christina and Ewer, Katie and Folegatti Pedro M. and Fowler, Jamie and Hill Adrian V. S. and Kerridge, Simon and Minassian Angela M. and Mongkolspaya, Juthathip and Farooq Mujadidi Yama, and Plested, Emma and Ramasamy Maheshi N. and Robinson, Hannah and Sanders, Helen and Sheehan, Emma and Smith, Holly and Snape Matthew D. and Song, Rinn and Woods, Danielle and Screaton Gavin R. and Gilbert Sarah C. and Voysey, Merryn and Pollard, Andrew and Lambe, Teresa and Group,, 2021. The Oxford COVID Vaccine, Tolerability and Immunogenicity After a Late Second Dose or a Third Dose of ChAdOx1 nCoV-19 (AZD1222)..

Hannah Ritchie, E.O.-O., Diana Beltekian, Edouard Mathieu, Joe Hasell, Bobbie Macdonald, Charlie Giattino, Cameron Appel and Max Roser, 2021. Coronavirus (COVID-19) Vaccinations, COVID-19 vaccine doses administered per 100 people, Apr 1, 2021. Our World in Data.

Iacobucci, G., 2021. Covid-19: Single vaccine dose is 33% effective against variant from India, data show. BMJ (Clinical research ed 373, 1346.

Jeewandara, C., Jayathilaka, D., Gomes, L., Wijewickrama, A., Narangoda, E., Idampitiya, D., Guruge, D., Wijayamuni, R., Manilgama, S., Ogg, G.S., Tan, C.W., Wang, L.F., Malavige, G.N., 2021. SARS-CoV-2 neutralizing antibodies in patients with varying severity of acute COVID-19 illness. Sci Rep 11, 2062.

Jeewandara, C., Kamaladasa, A., Pushpakumara, P.D., Jayathilaka, D., Abayrathna, I.S., Danasekara, S., Guruge, D., Ranasinghe, T., Dayarathne, S., Pathmanathan, T., Somathilaka, G., Madhusanka, D., Tanussiya, S., Tpj, T., Kuruppu, H., Wijesinghe, A., Thashmi, N., Milroy, D., Nandasena, A., Sanjeewani, N., Wijayamuni, R., Samaraweera, S., Schimanski, L., Tan, T.K., Dong, T., Ogg, G.S., Townsend, A., Malavige, G.N., 2021. Antibody and T cell responses to a single dose of the AZD1222/Covishield vaccine in previously SARS-CoV-2 infected and naïve health care workers in Sri Lanka. medRxiv, 2021.2004.2009.21255194.

Jeewandara, C., Ogg, G.S., Malavige, G.N., 2018. Cultured ELISpot Assay to Investigate Dengue Virus Specific T-Cell Responses. Methods Mol Biol 1808, 165–171.

Kalimuddin, S., Tham, C.Y.L., Qui, M., de Alwis, R., Sim, J.X.Y., Lim, J.M.E., Tan, H.C., Syenina, A., Zhang, S.L., Le Bert, N., Tan, A.T., Leong, Y.S., Yee, J.X., Ong, E.Z., Ooi, E.E., Bertoletti, A., Low, J.G., 2021. Early T cell and binding antibody responses are associated with COVID-19 RNA vaccine efficacy onset. Med (N Y) 2, 682–688 e684.

Kamaladasa, A., Gunasekara, B., Jeewandara, C., Jayathilaka, D., Wijewickrama, A., Guruge, D., Wijayamuni, R., t, K.T., Ogg, G.S., Townsend, A., Malavige, G.N., 2021. Comparison of two assays to detect IgG antibodies to the receptor binding domain of the SARSCoV2 as a surrogate marker for assessing neutralizing antibodies in COVID-19 patients. Int J Infect Dis.

Malavige, G.N., Jones, L., Kamaladasa, S.D., Wijewickrama, A., Seneviratne, S.L., Black, A.P., Ogg, G.S., 2008. Viral load, clinical disease severity and cellular immune responses in primary varicella zoster virus infection in Sri Lanka. PloS one 3, e3789.

Medicine, J.H.U.a., 2021. Coronavrus Resource Centre, CRITICAL TRENDS: TRACKING CRITICAL DATA. John Hopkins University.

Nasreen, S., Chung, H., He, S., Brown, K.A., Gubbay, J.B., Buchan, S.A., Fell, D.B., Austin, P.C., Schwartz, K.L., Sundaram, M.E., Calzavara, A., Chen, B., Tadrous, M., Wilson, K., Wilson, S.E., Kwong, J.C., Investigators, o.b.o.t.C.I.R.N.P.C.N., 2021. Effectiveness of COVID-19 vaccines against variants of concern in Ontario, Canada. medRxiv, 2021.2006.2028.21259420.

Nguyen-Contant, P., Embong, A.K., Kanagaiah, P., Chaves, F.A., Yang, H., Branche, A.R., Topham, D.J., Sangster, M.Y., 2020. S Protein-Reactive IgG and Memory B Cell Production after Human SARS-CoV-2 Infection Includes Broad Reactivity to the S2 Subunit. mBio 11.

Peng, Y., Mentzer, A.J., Liu, G., Yao, X., Yin, Z., Dong, D., Dejnirattisai, W., Rostron, T., Supasa, P., Liu, C., Lopez-Camacho, C., Slon-Campos, J., Zhao, Y., Stuart, D.I., Paesen, G.C., Grimes, J.M., Antson, A.A., Bayfield, O.W., Hawkins, D., Ker, D.S., Wang, B., Turtle, L., Subramaniam, K., Thomson, P., Zhang, P., Dold, C., Ratcliff, J., Simmonds, P., de Silva, T., Sopp, P., Wellington, D., Rajapaksa, U., Chen, Y.L., Salio, M., Napolitani, G., Paes, W., Borrow, P., Kessler, B.M., Fry, J.W., Schwabe, N.F., Semple, M.G., Baillie, J.K., Moore, S.C., Openshaw, P.J.M., Ansari, M.A., Dunachie, S., Barnes, E., Frater, J., Kerr, G., Goulder, P., Lockett, T., Levin, R., Zhang, Y., Jing, R., Ho, L.P., Oxford Immunology Network Covid-19 Response, T.c.C., Investigators, I.C., Cornall, R.J., Conlon, C.P., Klenerman, P., Screaton, G.R., Mongkolsapaya, J., McMichael, A., Knight, J.C., Ogg, G., Dong, T., 2020. Broad and strong memory CD4(+) and CD8(+) T cells induced by SARS-CoV-2 in UK convalescent individuals following COVID-19. Nature immunology 21, 1336–1345.

Pimenta, D., Yates, C., Pagel, C., Gurdasani, D., 2021. Delaying the second dose of covid-19 vaccines. BMJ (Clinical research ed 372, n710.

Ramasamy, M.N., Minassian, A.M., Ewer, K.J., Flaxman, A.L., Folegatti, P.M., Owens, D.R., Voysey, M., Aley, P.K., Angus, B., Babbage, G., Belij-Rammerstorfer, S., Berry, L., Bibi, S., Bittaye, M., Cathie, K., Chappell, H., Charlton, S., Cicconi, P., Clutterbuck, E.A., Colin-Jones, R., Dold, C., Emary, K.R.W., Fedosyuk, S., Fuskova, M., Gbesemete, D., Green, C., Hallis, B., Hou, M.M., Jenkin, D., Joe, C.C.D., Kelly, E.J., Kerridge, S., Lawrie, A.M., Lelliott, A., Lwin, M.N., Makinson, R., Marchevsky, N.G., Mujadidi, Y., Munro, A.P.S., Pacurar, M., Plested, E., Rand, J., Rawlinson, T., Rhead, S., Robinson, H., Ritchie, A.J., Ross-Russell, A.L., Saich, S., Singh, N., Smith, C.C., Snape, M.D., Song, R., Tarrant, R., Themistocleous, Y., Thomas, K.M., Villafana, T.L., Warren, S.C., Watson, M.E.E., Douglas, A.D., Hill, A.V.S., Lambe, T., Gilbert, S.C., Faust, S.N., Pollard, A.J., Oxford, C.V.T.G., 2021. Safety and immunogenicity of ChAdOx1 nCoV-19 vaccine administered in a prime-boost regimen in young and old adults (COV002): a single-blind, randomised, controlled, phase 2/3 trial. Lancet 396, 1979–1993.

Sadarangani, M., Marchant, A., Kollmann, T.R., 2021. Immunological mechanisms of vaccine-induced protection against COVID-19 in humans. Nature reviews. Immunology.

Tan, C.W., Chia, W.N., Qin, X., Liu, P., Chen, M.I., Tiu, C., Hu, Z., Chen, V.C., Young, B.E., Sia, W.R., Tan, Y.J., Foo, R., Yi, Y., Lye, D.C., Anderson, D.E., Wang, L.F., 2020. A SARS-CoV-2 surrogate virus neutralization test based on antibody-mediated blockage of ACE2-spike protein-protein interaction. Nature biotechnology 38, 1073–1078.

Tauh. T M.M., Meyler, P., Lee, S.M., 2021. An updated look at the 16-week window between doses of vaccines in BC for COVID-19. BC Medical Journal 63, 102–103.

Townsend, A., Rijal, P., Xiao, J., Tan, T.K., Huang, K.-Y.A., Schimanski, L., Huo, J., Gupta, N., Rahikainen, R., Matthews, P.C., Crook, D., Hoosdally, S., Street, T., Rudkin, J., Stoesser, N., Karpe, F., Neville, M., Ploeg, R., Oliveira, M., Roberts, D.J., Lamikanra, A.A., Tsang, H.P., Bown, A., Vipond, R., Mentzer, A.J., Knight, J.C., Kwok, A., Screaton, G., Mongkolsapaya, J., Dejnirattisai, W., Supasa, P., Klenerman, P., Dold, C., Baillie, K., Moore, S.C., Openshaw, P.J., Semple, M.G., Turtle, L.C., Ainsworth, M., Allcock, A., Beer, S., Bibi, S., Clutterbuck, E., Espinosa, A., Mendoza, M., Georgiou, D., Lockett, T., Martinez, J., Perez, E., Sanchez, V., Scozzafava, G., Sobrinodiaz, A., Thraves, H., Joly, E., 2020. A haemagglutination test for rapid detection of antibodies to SARS-CoV-2. Nature Communications, 2020.2010.2002.20205831.

Turner, J.S., Kim, W., Kalaidina, E., Goss, C.W., Rauseo, A.M., Schmitz, A.J., Hansen, L., Haile, A., Klebert, M.K., Pusic, I., O’Halloran, J.A., Presti, R.M., Ellebedy, A.H., 2021. SARS-CoV-2 infection induces long-lived bone marrow plasma cells in humans. Nature 595, 421–425.

Voysey, M., Clemens, S.A.C., Madhi, S.A., Weckx, L.Y., Folegatti, P.M., Aley, P.K., Angus, B., Baillie, V.L., Barnabas, S.L., Bhorat, Q.E., Bibi, S., Briner, C., Cicconi, P., Collins, A.M., Colin-Jones, R., Cutland, C.L., Darton, T.C., Dheda, K., Duncan, C.J.A., Emary, K.R.W., Ewer, K.J., Fairlie, L., Faust, S.N., Feng, S., Ferreira, D.M., Finn, A., Goodman, A.L., Green, C.M., Green, C.A., Heath, P.T., Hill, C., Hill, H., Hirsch, I., Hodgson, S.H.C., Izu, A., Jackson, S., Jenkin, D., Joe, C.C.D., Kerridge, S., Koen, A., Kwatra, G., Lazarus, R., Lawrie, A.M., Lelliott, A., Libri, V., Lillie, P.J., Mallory, R., Mendes, A.V.A., Milan, E.P., Minassian, A.M., McGregor, A., Morrison, H., Mujadidi, Y.F., Nana, A., O’Reilly, P.J., Padayachee, S.D., Pittella, A., Plested, E., Pollock, K.M., Ramasamy, M.N., Rhead, S., Schwarzbold, A.V., Singh, N., Smith, A., Song, R., Snape, M.D., Sprinz, E., Sutherland, R.K., Tarrant, R., Thomson, E.C., Torok, M.E., Toshner, M., Turner, D.P.J., Vekemans, J., Villafana, T.L., Watson, M.E.E., Williams, C.J., Douglas, A.D., Hill, A.V.S., Lambe, T., Gilbert, S.C., Pollard, A.J., Oxford, C.V.T.G., 2021a. Safety and efficacy of the ChAdOx1 nCoV-19 vaccine (AZD1222) against SARS-CoV-2: an interim analysis of four randomised controlled trials in Brazil, South Africa, and the UK. Lancet 397, 99–111.

Voysey, M., Costa Clemens, S.A., Madhi, S.A., Weckx, L.Y., Folegatti, P.M., Aley, P.K., Angus, B., Baillie, V.L., Barnabas, S.L., Bhorat, Q.E., Bibi, S., Briner, C., Cicconi, P., Clutterbuck, E.A., Collins, A.M., Cutland, C.L., Darton, T.C., Dheda, K., Dold, C., Duncan, C.J.A., Emary, K.R.W., Ewer, K.J., Flaxman, A., Fairlie, L., Faust, S.N., Feng, S., Ferreira, D.M., Finn, A., Galiza, E., Goodman, A.L., Green, C.M., Green, C.A., Greenland, M., Hill, C., Hill, H.C., Hirsch, I., Izu, A., Jenkin, D., Joe, C.C.D., Kerridge, S., Koen, A., Kwatra, G., Lazarus, R., Libri, V., Lillie, P.J., Marchevsky, N.G., Marshall, R.P., Mendes, A.V.A., Milan, E.P., Minassian, A.M., McGregor, A., Mujadidi, Y.F., Nana, A., Padayachee, S.D., Phillips, D.J., Pittella, A., Plested, E., Pollock, K.M., Ramasamy, M.N., Ritchie, A.J., Robinson, H., Schwarzbold, A.V., Smith, A., Song, R., Snape, M.D., Sprinz, E., Sutherland, R.K., Thomson, E.C., Torok, M.E., Toshner, M., Turner, D.P.J., Vekemans, J., Villafana, T.L., White, T., Williams, C.J., Douglas, A.D., Hill, A.V.S., Lambe, T., Gilbert, S.C., Pollard, A.J., Oxford, C.V.T.G., 2021b. Single-dose administration and the influence of the timing of the booster dose on immunogenicity and efficacy of ChAdOx1 nCoV-19 (AZD1222) vaccine: a pooled analysis of four randomised trials. Lancet 397, 881–891.

Wijeratne, D.T., Fernando, S., Gomes, L., Jeewandara, C., Jayarathna, G., Perera, Y., Wickramanayake, S., Wijewickrama, A., Ogg, G.S., Malavige, G.N., 2019. Association of dengue virus-specific polyfunctional T-cell responses with clinical disease severity in acute dengue infection. Immun Inflamm Dis.

Woldemeskel, B.A., Garliss, C.C., Blankson, J.N., 2021. SARS-CoV-2 mRNA vaccines induce broad CD4+ T cell responses that recognize SARS-CoV-2 variants and HCoV-NL63. The Journal of clinical investigation 131.

