## Supplementary figures and images for "Immune responses to a single dose of the AZD1222/Covishield vaccine at 16 weeks in individuals in Sri Lanka"

### Supplementary figures

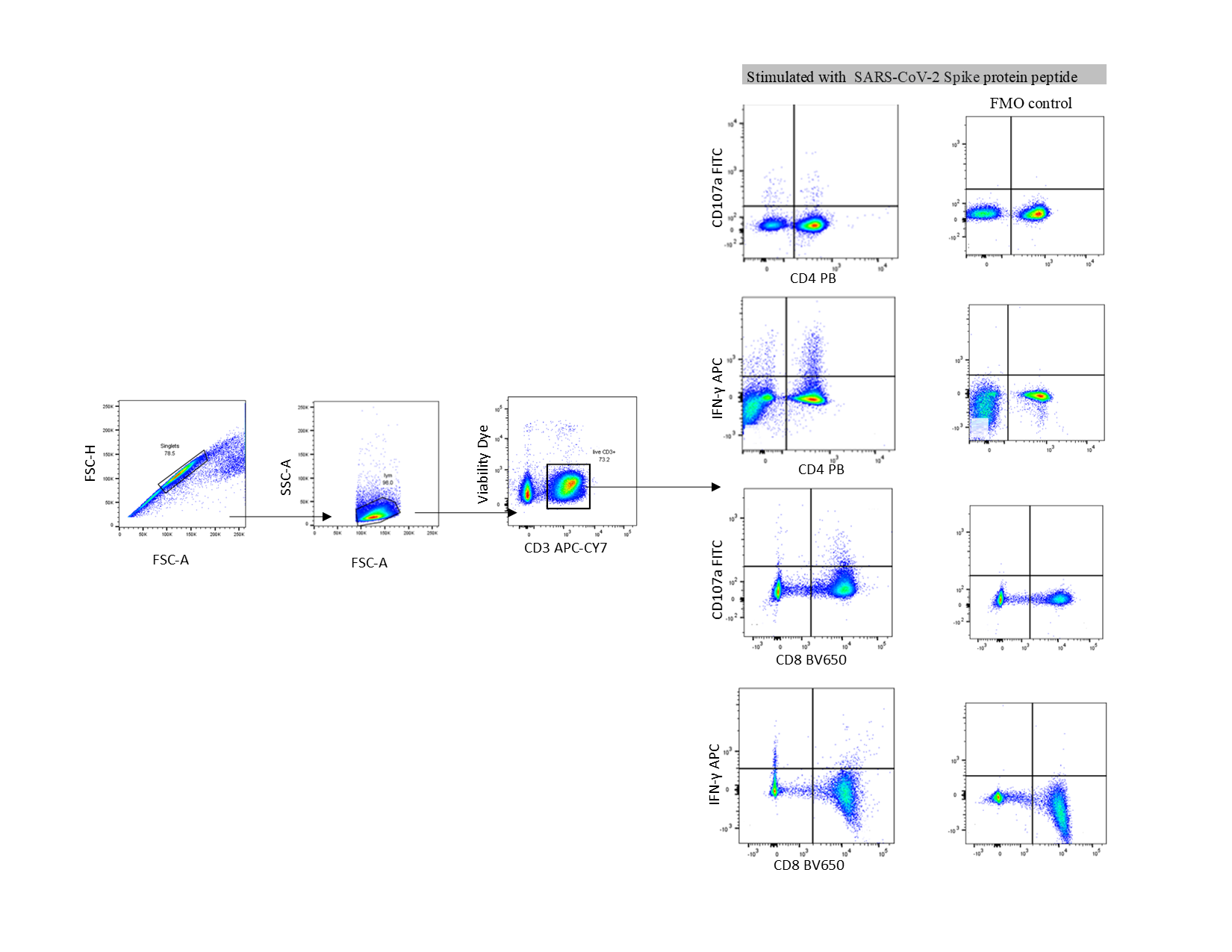
